# Infection, inflammation and post-stroke cognitive impairment

**DOI:** 10.1101/2023.07.19.23292862

**Authors:** Elise Milosevich, Nele Demeyere, Sarah T. Pendlebury

**Affiliations:** Department of Experimental Psychology, University of Oxford, UK; Wolfson Centre for Prevention of Stroke and Dementia, Wolfson Building, Nuffield Department of Clinical Neurosciences, University of Oxford, UK; NIHR Oxford Biomedical Research Centre and Departments of General Medicine and Geratology, John Radcliffe Hospital, Oxford, UK

**Keywords:** Stroke, Infection, Inflammation, Cognition, Cognitive dysfunction

## Abstract

**Background:** Infection and inflammation are risk factors for dementia in population-based cohorts, however studies in stroke are scarce. We determined the prevalence of stroke-associated infection and routinely measured inflammatory biomarkers during hospitalization for stroke, and their associations with global and domain-specific cognition acutely and at 6-months.

**Methods:** A prospective stroke cohort completed the Oxford Cognitive Screen ≤2 weeks and at 6 months. Global post-stroke cognitive impairment (PSCI) was defined as proportion of domains impaired and domain-specific impairment was binarized as present/absent. Infection, inflammatory markers (C-reactive protein-CRP, white cell count-WCC, and neutrophil/lymphocyte ratio-NLR), and systemic inflammatory response syndrome-SIRS, were ascertained throughout admission through linkage to electronic patient records-EPRs supplemented by hand-searches. Associations of infection, inflammatory markers, and SIRS, with acute and 6-month global and domain-specific cognitive impairment were analyzed using multivariable regression adjusting for demographic/vascular factors and stroke severity.

**Results:** Among 255 patients (mean/SD age=73.9/12.6 years, 46.3% female, mean/SD education=12.6/3.7 years, median NIHSS=5, range 0–30), infection was present in 90 (35.3%) patients at mean/SD 4.4/6.9 days post-stroke, most commonly pneumonia (47/90, 52%) and urinary tract infection (39/90, 43%). Admission WCC was elevated in 64 (25.1%, mean/SD=9.5/3.2×10^9^/L), CRP in 105 (41.2%, mean/SD=27.5/50.9 mg/L), NLR in 97 (55.7%, mean/SD=5.5/4.5) and SIRS in 53 (26.6% with 45.2% positive overall during admission). Infection was associated with acute and 6-month PSCI (*p*<0.05_adj_) with stronger associations acutely for severe infection (infection+SIRS,p=0.02_adj_). Acute impairments in language, executive function and attention domains and 6-month impairment in number processing (all *p*<0.05_adj_) were associated with infection. No significant relationships were found for any biomarker and cognitive impairment.

**Conclusion:** Infection and elevations in routinely measured inflammatory biomarkers are common after stroke, however only infection is a risk factor for PSCI, suggesting that the rise in these biomarkers may be non-specific. Infection may present a tractable target for reducing PSCI.

## INTRODUCTION

Despite modest improvement in stroke outcomes^1^, post-stroke cognitive impairment (PSCI) and dementia remain highly prevalent and often disabling.^2, 3^ PSCI risk is mediated in part by pre-existing brain susceptibility^4^ and the impact of the stroke lesion. However, these factors do not explain all the variance in risk.^2, 5^ Within non-stroke populations, there is emerging evidence linking infection to future development of all cause dementia and in particular, vascular dementia^6, 7^ with a dose response effect and preliminary data suggest that infection may also be important in PSCI.^8, 9^

Infection is the most frequent complication of stroke, affecting ∼30% of patients, with pneumonia and urinary tract infections (UTI) being most common.^10^ Infections are associated with increased stroke morbidity and mortality.^11^ Stroke Unit care may result in better outcomes in part through preventing such complications,^12^ however there are few data specifically on cognitive function. In one “Big Data” study, infection after stroke including during the follow-up period after discharge was associated with a >40% increase in the risk of early dementia with greater impact of more severe (hospital) infection (3-12 months post-stroke).^8^ However, dementia was identified solely through administrative diagnostic coding, which is known to have poor sensitivity.^13,14^ Infections may act through systemic inflammatory pathways to trigger a disordered microglial response in the aging brain resulting in progression of Alzheimer’s disease (AD)^15^ and small vessel disease (SVD), via vascular inflammation, endothelial dysfunction and BBB disruption.^16^

Stroke-associated biomarkers of inflammation may also be associated with PSCI,^17–19^ including C-reactive protein (CRP),^19^ neutrophil-lymphocyte ratio (NLR),^17^ erythrocyte sedimentation rate (ESR),^20^ terminal C5b-9 complement complex (TCC), IL-6, and MIP-1α.^21^ Ischaemia leads to the induction of proteases and free radicals, blood brain barrier (BBB) opening, and ultimately myelin breakdown and oligodendrocyte death.^22^ A more proinflammatory immune signature in peripheral blood at 2 days post-stroke has been linked to PSCI within the first year^23^ and acute inflammation appears more predictive than chronic inflammation for post-stroke cognitive outcome at 36 months.^21^

However, there are few studies that have examined both stroke-associated infection and biomarkers of inflammation in relation to PSCI despite the fact that inflammation would be expected to co-occur with infection. It is important for understanding both mechanisms and risk prediction whether infection or acute inflammation from whichever cause (or both) increase the risk of PSCI and to what extent these are independent of confounding factors. Additionally, given that infection appears to predispose individuals predominantly to vascular dementia, one might expect a greater impact on those with a vascular cognitive profile^24^ though there are few data particularly in relation to PSCI.

Therefore, this study in acute stroke patients sought to determine (i) the prevalence, subtype and severity of infection, and the prevalence of elevated routinely collected markers of inflammation during hospitalization, as well as (ii) the relationship between acute infection, inflammatory markers, and global and domain-specific cognitive function acutely and at 6 months follow-up.

## METHODS

A consecutive sample of acute stroke patients (n=866) was recruited through the Oxford Cognitive Screening-OCS programme^25^ (2012-2019, National Research Ethics Committee-REC 14/LO/0648; 18/SC/0550) based within the regional acute stroke unit, John Radcliffe Hospital, UK. Manually collected research data were supplemented by routinely acquired electronic patient record (EPR) data through linkage to the Oxford Cognitive Comorbidity, Frailty, and Ageing Research database (ORCHARD, REC reference 18/SC/0184). For the present study, consecutive patients recruited January 2015 to September 2019 with both acute and 6-month follow-up data were included, as EPR, and therefore ORCHARD, commenced in 2015 (Figure 1). Acute stroke patients were included if aged 18 years or older, able to concentrate for approximately 20 minutes as judged by the multidisciplinary team and had sufficient English language comprehension to understand assessment instructions. All participants included provided written or witnessed informed consent and STROBE reporting guidance for cohort studies was followed.

**Figure 1.**
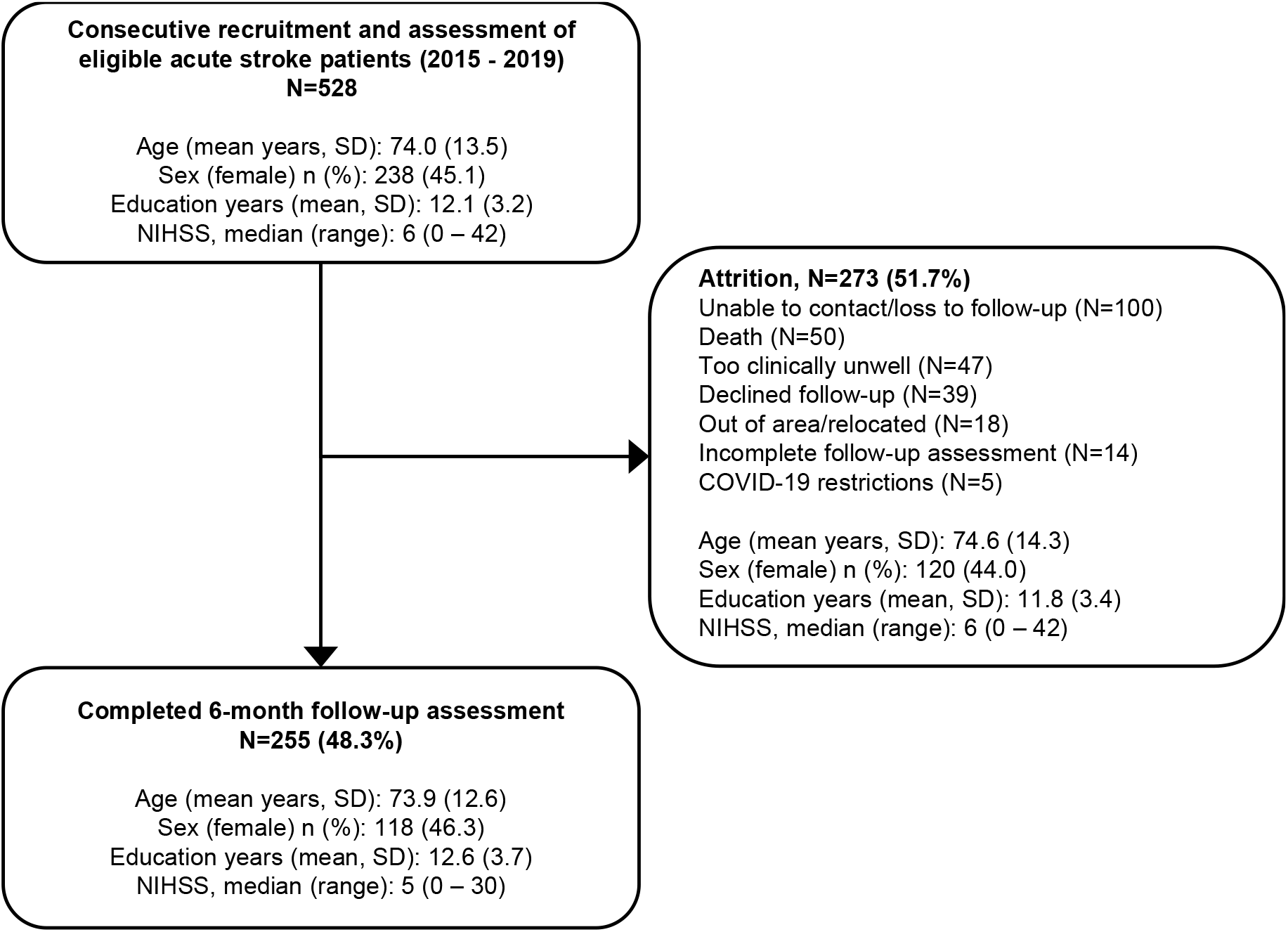
Flowchart of study cohort.

### Participants

All included participants received cognitive screening with the OCS within 2 weeks of stroke and again at 6-month follow-up. Data collection also included patient demographics, comorbidities, vascular risk factors and stroke characteristics, including stroke severity evaluated using the National Institutes of Health Stroke Scale (NIHSS).^26^ Diagnosis of infection, routinely acquired laboratory test results, Charlson Comorbidity Index (CCI)^27^ and secondary events (e.g., hydrocephalus) were obtained through linkage to ORCHARD and hand-searching of electronic medical records.^28^ (Table S1).

### Infections and inflammatory markers

Infections were categorized into urinary tract infection (UTI), pneumonia, other respiratory, gastrointestinal, hepatitis, soft tissue, central nervous system, blood, and other (e.g., sinusitis, oesophageal candidiasis). Suspected UTI was defined as symptomatic with positive dipstick for leukocyte esterase and/or nitrate, and received treatment, while confirmed UTI required symptoms, a positive culture, and received appropriate therapy.^29^ Pneumonia was binarized as suspected (symptoms, clinical diagnosis, treated) or confirmed (chest radiograph). All remaining infections were confirmed with positive signs/symptoms and laboratory tests (e.g., positive culture or antigen detection). The presence of a hospital discharge ICD-10 diagnosis of sepsis was recorded.^30^

Severity of infection inflammatory response was defined by the systemic inflammatory response syndrome (SIRS) criteria^31^ (≥2 of: heart rate >90/min; respiratory rate >20/min; temperature <36°C or >38°C; or white cell count (WCC) <4×10^9^/L or >12×10^9^/L. Total days (consecutive or non-consecutive) SIRS-positive during admission period was calculated. Routinely acquired inflammatory markers, including CRP, WCC, lymphocyte count, and neutrophil count, were recorded from admission and any repeat sampling during admission was reviewed to identify peak values of CRP and WCC. Admission neutrophil count divided by the lymphocyte count yielded the Neutrophil-Lymphocyte ratio (NLR) with normal reference range for adults of 0.78-3.53.^32^ Reference intervals employed were derived from patient recruitment site (John Radcliffe Hospital, Oxford University Hospitals NHS Foundation Trust).

### Cognitive assessment

Cognition was assessed within 2 weeks post-stroke and at 6 months using the Oxford Cognitive Screen (OCS).^25^ The OCS covers a broad range of cognitive domains and was designed specifically for use in acute stroke taking 15-20 minutes to complete (Table S2). All 12 subtest scores were categorized into 6 cognitive domains: language (picture naming, semantic understanding, sentence reading), attention (egocentric attention/sustained attention, allocentric attention), executive function (trail-making), memory (orientation, verbal memory, episodic memory), praxis (gesture imitation), and number processing (calculation and number writing). Subtests were binarized into impaired or unimpaired based on normative scores for each subtest (cut-offs were set at 5^th^ centile).^25^ Severity of global cognitive impairment was characterized by the proportion of subtests impaired across all domains. Domain-specific impairment was defined as at least one subtest impaired within a domain. The OCS was administered by trained psychologists and occupational therapists at the bedside acutely and at in-person follow-up.

### Statistical analysis

Descriptive statistics were used to summarize sample demographics, clinical characteristics and the prevalence of post-stroke infection, SIRS, and elevated inflammatory markers (Aim 1). Differences in clinical characteristics and cognitive function between patients with and without infection were determined using *t*-test and Fisher’s exact test as appropriate. To address Aim 2, first, univariate linear regression analyses were used to assess associations between stroke-associated infection, SIRS, and inflammatory markers (CRP, NLR, WCC), and severity of global cognitive impairment acutely and at 6 months. Next, if a significant univariate relationship was present, multivariate regression was performed adjusted for available variables known to be associated with cognitive impairment after stroke: age, sex, years of education, stroke severity (NIHSS), previous stroke, atrial fibrillation, hypertension, diabetes, and smoking.^33–35^ Multivariate regression was also conducted to examine the relationship between severe infection (infection+SIRS) and cognitive impairment acutely and at 6 months. Lastly, logistic regression was applied to investigate the relationship between post-stroke infection/inflammation and binarized domain-specific cognitive function (impaired/unimpaired) acutely and at 6 months. Each of these models again adjusted for demographics, stroke-related and vascular risk factors. Missing NIHSS data were addressed through multiple imputation. Sensitivity analyses were undertaken restricted to participants with complete NIHSS data (findings were broadly similar; see supplemental materials). For all analyses, *p* values <0.05 were considered statistically significant. Unstandardized beta (*B*), standard error for the unstandardized beta (*SE*), the *t* statistic and the probability value (*p*-value) were reported for all regression analyses. Odds ratios and 95% confidence intervals were presented for logistic regression analyses. All analyses were carried out in R version 4.1.3.^36^

## RESULTS

Among 255 stroke survivors, (mean/SD age 73.9/12.6 years, 118 (46.3%) female, mean/SD years of education 12.6/3.7, 120 (47.1%) with NIHSS >5) cognitive assessment was completed at a mean/SD 3.6/3.8 days post-stroke, with a mean/SD follow-up interval of 6.7/1.1 months (Figure 1; Table 1). Acutely, 213 (83.5%) were impaired on at least one subtest in a cognitive domain, with multi-domain impairments being most common (n=166 (65.1%)). At 6 months, 192 (75.3%) were impaired and 175 (49.8%) demonstrated multi-domain impairments. Domain-specific impairments ranged from 26.9% in praxis to 44.9% in attention acutely, and 17.9% in number processing to 35.9% in attention at follow-up (Table S2).

**Table 1.** Cohort characteristics.

|  | All patients<br>N=255 | No Infection<br>N=165 | Infection (≥1)<br>N=90 | p value |
| --- | --- | --- | --- | --- |
| Age, mean years/SD | 73.9/12.6 | 72.7/13.2 | 76.3/11.0 | 0.03 |
| Sex, female, N (%) | 118 (46.3) | 74 (44.9) | 44 (48.9) | 0.54 |
| Education years, mean/SD | 12.6/3.7 | 12.79/3.7 | 12.2/3.6 | 0.25 |
| *Stroke subtype*, N (%) |  |  |  |  |
| Ischemic | 211 (82.8) | 138 (83.6) | 73 (81.1) | 0.61 |
| Haemorrhagic | 42 (16.5) | 25 (15.2) | 17 (18.9) | 0.48 |
| Mixed | 2 (78.4) | 2 (1.2) | 0 (0.0) | 0.54 |
| First-ever stroke, N (%) | 178 (69.8) | 117 (70.9) | 61 (67.8) | 0.67 |
| NIHSS,<sup>1</sup> median (range) | 5 (0 – 30) | 4 (0 – 24) | 6 (0–30) | 0.16 |
| NIHSS<5, N (%) | 95 (44.2) | 73 (51.0) | 22 (15.4) | <0.01 |
| NIHSS≥5, N (%) | 120 (55.8) | 70 (49.0) | 50 (69.4) |  |
| Independent at admission,<sup>2</sup> N (%) | 232 (91.0) | 158 (95.8) | 74 (82.2) | <0.01 |
| Charlson Comorbidity Index score, median (range) | 1 (0–6) | 1 (0–6) | 1 (0–5) | 0.11 |
| Atrial fibrillation, N (%) | 61 (23.9) | 36 (21.8) | 25 (27.8) | 0.28 |
| Hypertension, N (%) | 146 (57.3) | 89 (53.9) | 57 (63.3) | 0.19 |
| Diabetes mellitus, N (%) | 46 (18.0) | 31 (18.8) | 15 (16.7) | 0.74 |
| *Smoking*, N (%) |  |  |  |  |
| Current | 20 (7.8) | 14 (8.5) | 6 (6.7) | 0.81 |
| Past | 26 (10.2) | 15 (9.1) | 11 (12.2) | 0.52 |
| Never | 209 (82.0) | 136 (82.4) | 73 (81.1) | 0.87 |
| Hospital stay, days mean/SD | 11.2/10.4 | 8.0/6.7 | 17.0/13.1 | <0.01 |
| T1 assessment days post-stroke, mean/SD | 3.6/3.8 | 3.2/3.3 | 4.4/4.6 | 0.02 |
| Acute cognitive impairment (OCS) | 213 (83.5) | 129 (78.2) | 84 (93.3) | <0.01 |
| Single-domain impairment, N (%) | 47 (18.4) | 33 (20.0) | 14 (15.6) | 0.40 |
| Multi-domain impairment, N (%) | 166 (65.1) | 96 (58.2) | 70 (77.8) | <0.01 |
| T2 assessment months post-stroke, mean/SD | 6.7/1.1 | 6.7/1.0 | 6.8/1.3 | 0.59 |
| 6-month cognitive impairment (OCS) | 201 (78.8) | 124 (75.2) | 77 (86.7) | 0.06 |
| Single-domain impairment, N (%) | 64 (25.1) | 46 (27.9) | 18 (20.0) | 0.18 |
| Multi-domain impairment, N (%) | 137 (53.7) | 78 (60.6) | 59 (65.6) | <0.01 |

CCI: Charlson Comorbidity Index; NIHSS: National Institute of Health Stroke Scale.
<sup>1</sup>NIHSS data available for n=215 (84.3%); n=143 no infection, n=72 infection.<sup>2</sup>Independence was categorized by not receiving any support (required family/carer).

### Prevalence of elevated inflammatory markers and systemic inflammatory response syndrome (SIRS) acutely after stroke

On admission, the most frequently elevated biomarker was NLR in 142 (55.7%) patients, followed by CRP in 105 (41.2%), and WCC in 64 (25.1%) (Table 2). Peak CRP (mean/SD=52.0/74.7 mg/L) occurred most often on day 3 (median; range 0-52) and was elevated at some point in the majority (n=137; 72.1%). SIRS-positive on admission occurred in 53/199 (26.6%, in 56 patients SIRS was unavailable) (Table 3), and 90 (45.2%) were SIRS-positive at least once during hospitalization (with a mean/SD of 2.7/3.3 days; range 1-26) days. Sepsis was identified in 8 (3.1%) patients during hospitalization.

**Table 2.** Post-stroke inflammatory biomarkers during hospital admission.

| Inflammatory biomarker | Mean/SD | Median (range) | Outside reference interval<br>N (%) |
| --- | --- | --- | --- |
| WCC (on admission) | 9.5/3.2 | 9.0 (4.2–20.6) | 64 (25.1) |
| WCC (peak) | 10.8/4.1 | 10.0 (4.2–38.0) | 101 (39.6) |
| Day of peak WCC | 2.5/5.1 | 0 (0–33) |  |
| CRP* (on admission) | 28.5/50.9 | 7.6 (0.4–282.9) | 105 (41.2) |
| CRP* (peak) | 52.0/74.7 | 18.4 (0.5–461.5) | 137 (72.1) |
| Day of peak CRP | 5.1/7.3 | 3 (0–52) |  |
| Lymphocyte count (on admission) | 1.7/0.9 | 1.5 (0.3–7.6) | 5 (2.0) |
| Neutrophil count (on admission) | 6.9/3.1 | 6.1 (2.1–17.3) | 97 (38.0) |
| NLR (on admission) | 5.5/4.5 | 3.8 (0.7–27.1) | 142 (55.7) |

WCC: White cell count, blood; CRP: C-reactive protein, plasma; NLR: Neutrophil-Lymphocyte ratio.
Reference intervals: WCC (4.0–11.0x10<sup>9</sup>/L); CRP (0–5 mg/L); Lymphocyte count 1.0–4.0x10<sup>9</sup>/L; Neutrophil count (2.0–7.0x10<sup>9</sup>/L); NLR (0.78–3.53); SBP (90–139 mmHg).
\*CRP data was available for 177/255 (69.4%) on admission and 190/255 (74.5%) patients throughout hospitalization.

**Table 3.** Prevalence of post-stroke infections, secondary events, and clinical features among all patients during admission period.

|  | N (%) | Days post-stroke confirmed |  |
| --- | --- | --- | --- |
|  |  | Mean/SD | Median (range) |
| Any infection | 90 (35.3) | 4.4/6.9 | 1.5 (0–49) |
| Single infection | 68 (26.7) |  |  |
| Multiple infections (>1) | 22 (8.6) |  |  |
| Pneumonia<sup>1</sup> | 47 (18.4) | 5.8/10.0 | 1.5 (0–49) |
| Urinary tract infection<sup>2</sup> | 39 (15.3) | 6.8/7.8 | 5 (0–42) |
| Soft tissue | 9 (3.5) | 6.6/7.4 | 1 (0–17) |
| Other respiratory | 8 (3.1) | 2.5/3.6 | 0.5 (0–9) |
| Blood | 8 (3.1) | 8.4/16.3 | 1.5 (0–48) |
| Other infection<sup>3</sup> | 5 (2.0) | 3.4/3.7 | 4 (0–9) |
| Gastrointestinal | 3 (1.2) | 21.0/23.8 | 12 (3–48) |
| Hepatitis (B) | 1 (0.4) | 8.0/NA | 8 (NA) |
| Sepsis | 8 (3.1) | 2.6/3.7 | 1 (0–9) |
| SIRS<sup>4</sup> positive on admission | 53/199 (26.6) |  |  |
| SIRS positive anytime during admission | 90/199 (45.2) |  |  |
| Total non-consecutive days positive |  | 2.7/3.3 | 2 (1–26) |
| *Oromotor/speech dysfunction* |  |  |  |
| Dysphagia | 103 (40.4) |  |  |
| Dysarthria/anarthria | 115 (45.1) |  |  |
| Nasogastric tube fed | 43 (16.9) |  |  |
| *Urinary dysfunction* |  |  |  |
| Catheterization | 22 (8.6) |  |  |
| Incontinence | 27 (10.6) |  |  |

Days post-stroke refers to first confirmed infection in instances of multiple. SIRS: Systemic Inflammatory Response Syndrome.
<sup>1</sup>Pneumonia confirmed: n=43(16.9%); suspected (clinical diagnosis, treated): n=4(0.4%).
<sup>2</sup>UTI confirmed: n=38(14.9%); suspected (clinical diagnosis, treated): n=1(1.6%).
<sup>3</sup>Other infection (one each): conjunctivitis, esophageal candidiasis, oral candidiasis, pyrexia of unknown origin, sinusitis.
<sup>4</sup>SIRS data available for n=199/255 patients.

### Infection and severity of cognitive impairment at acute and 6 months

The presence of any infection during hospitalization was associated with increased severity of global cognitive impairment acutely (*p*=0.005) and at 6 months (*p*<0.001) including after adjustment for confounders (p=0.037_adj_ and p=0,014_adj_) (Table 4; Table S5). Regarding different subtypes of infection, associations were seen for pneumonia both acutely (*p*<0.018_adj_) and at 6 months (*p*=0.031_adj_) but not for UTI in adjusted analyses (Table 4, Table S6). Severe infection (SIRS-positive) was associated with acute cognitive impairment (p=0.005_unad_j, *p*=0.023_adj_) but associations at 6-months attenuated after adjustment (p=0.008_unad_j, p=0.079_adj_) (Table 4). Conversely, infection with a SIRS-negative status was not associated with severity of cognitive impairment at either time point (Table 4, Table S7).

**Table 4.** Associations between post-stroke infection, inflammatory biomarkers, and severity of cognitive impairment acutely and at 6 months.

| *Unadjusted models* | Severity of cognitive impairment |  |  |  |  |  |  |  |
| --- | --- | --- | --- | --- | --- | --- | --- | --- |
| | Acute ($\leq 2$ weeks) | | | | 6 months | | | |
| | $\beta$ | SE | t | p | $\beta$ | SE | t | p |
| Infection (any) | 8.99 | 3.18 | 2.82 | **0.005** | 8.57 | 2.26 | 3.80 | **<0.001** |
| Pneumonia | 13.76 | 3.89 | 3.54 | **<0.001** | 9.03 | 3.27 | 2.76 | **0.006** |
| UTI | 5.57 | 4.28 | 1.30 | 0.195 | 6.27 | 3.09 | 2.03 | **0.043** |
| SIRS (admission) | 9.08 | 3.70 | 2.45 | **0.015** | 1.25 | 2.64 | 0.47 | 0.637 |
| SIRS (any time) | 5.87 | 3.31 | 1.77 | 0.078 | 2.36 | 2.34 | 1.01 | 0.315 |
| Infection, SIRS-neg | 5.36 | 5.14 | 1.04 | 0.298 | 6.47 | 3.59 | 1.80 | 0.073 |
| Infection, SIRS-pos | 10.49 | 3.91 | 2.68 | **0.008** | 7.70 | 2.74 | 2.82 | **0.005** |
| CRP (admission) | -0.01 | 0.04 | -0.17 | 0.863 | 0.01 | 0.03 | 0.39 | 0.696 |
| CRP (peak) | 0.01 | 0.02 | 0.54 | 0.590 | -0.02 | 0.02 | -0.95 | 0.341 |
| WCC (admission) | -0.21 | 0.49 | -0.42 | 0.672 | -0.36 | 0.35 | -1.03 | 0.302 |
| WCC (peak) | 0.61 | 0.38 | 1.60 | 0.112 | -0.02 | 0.27 | -0.06 | 0.956 |
| NLR (admission) | -0.48 | 0.34 | -1.39 | 0.165 | -0.21 | 0.25 | -0.86 | 0.392 |
| *Adjusted models* | $\beta$ | SE | t | p | $\beta$ | SE | t | p |
| Infection (any) | 6.35 | 3.02 | 2.10 | **0.037** | 6.13 | 2.47 | 2.49 | **0.014** |
| Pneumonia | 8.98 | 3.77 | 2.38 | **0.018** | 5.84 | 2.69 | 2.17 | **0.031** |
| UTI | 1.91 | 4.08 | 0.47 | 0.640 | 3.28 | 2.91 | 1.13 | 0.261 |
| SIRS (admission) | 9.19 | 3.63 | 2.54 | **0.012** | 1.80 | 2.38 | 0.75 | 0.452 |
| Infection, SIRS-neg | 4.11 | 4.99 | 0.82 | 0.411 | 3.92 | 4.24 | 0.80 | 0.425 |
| Infection, SIRS-pos | 8.84 | 3.85 | 2.30 | **0.023** | 5.09 | 2.88 | 1.76 | 0.079 |

Variables significant in univariate analyses were adjusted for demographics and vascular risk factors (age, sex, education, stroke severity, previous stroke, atrial fibrillation, hypertension, diabetes, and smoking). Severity of cognitive impairment was defined by proportion of Oxford Cognitive Screen tasks impaired acutely and at 6 months. SIRS-positive or negative refers to any time during admission period. Significance $p < 0.05$ .

### Inflammatory biomarkers, and severity of cognitive impairment at acute and 6 months

Univariate analyses showed no significant relationships between any inflammatory biomarker (CRP, WCC, or NLR) and cognitive impairment acutely or at 6 months (Table 4). In contrast, SIRS-positivity on admission was associated with cognitive impairment acutely (*p*=0.015) including after adjustment (*p*=0.012_adj_) (Table 4; Table S4), but not at 6 months (*p*=0.637_adj_).

### Infection and domain-specific cognitive impairment acutely and at 6 months

Stroke-associated infection was associated with domain-specific impairments in language (odds ratio [OR]_adj_ 1.725, 95% confidence interval [CI] 1.002–2.971, *p*<0.05), executive function (OR_adj_ 1.881, CI 1.050–3.367, *p*<0.05), and attention acutely (OR_adj_ 2.006, CI 1.142–3.522 *p*<0.05), and number processing (OR_adj_ 2.137, CI 1.073–4.254, *p*<0.05) at 6 months (Figure 2, Table S8).

**Figure 2.**
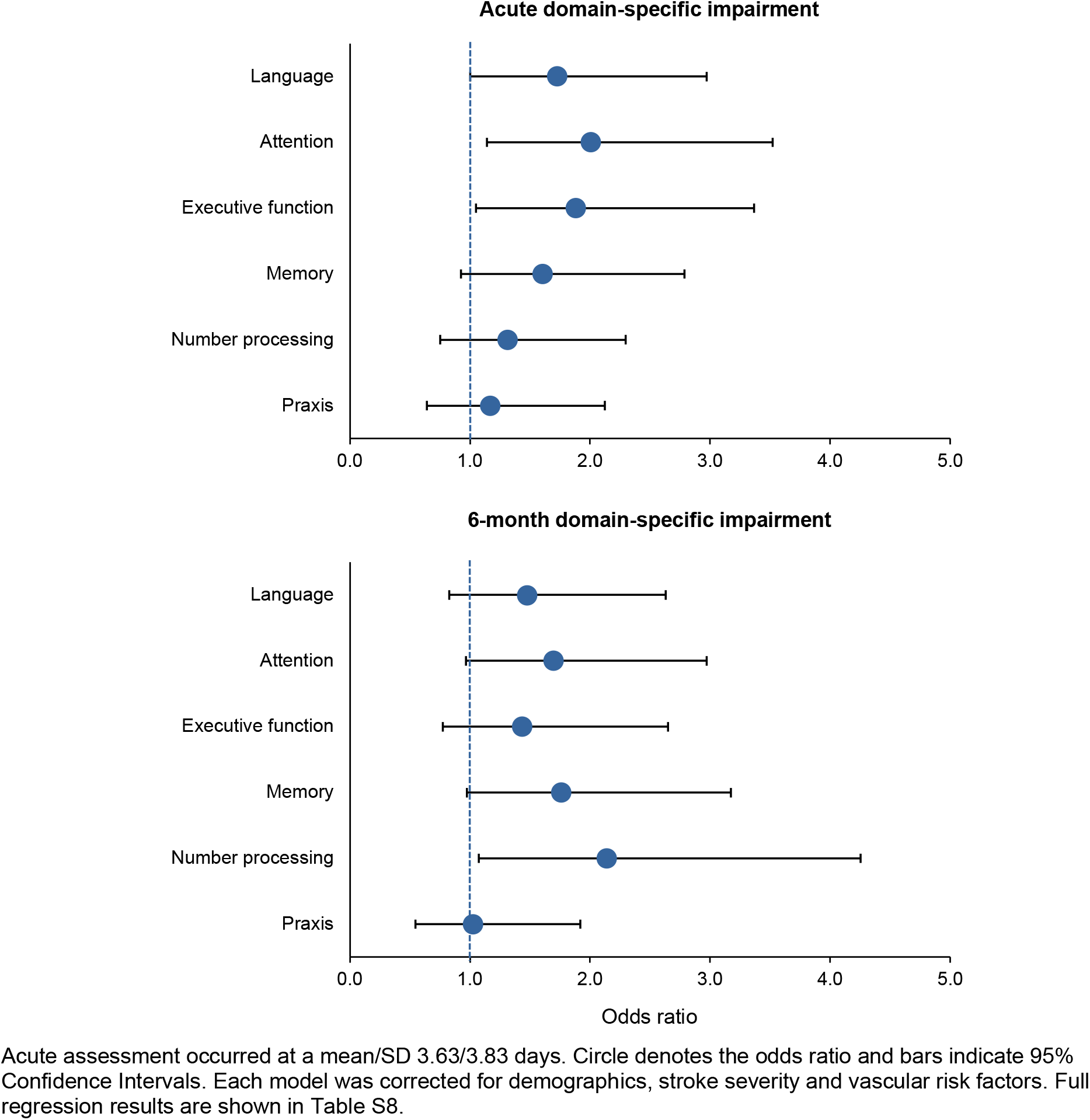
Odds ratios for the effect of acute post-stroke infection on domain-specific impairments acutely and at 6 months.

## DISCUSSION

Around one-third of this sample of acute stroke patients had at least one infection during hospitalization with pneumonia being most common, followed by UTI. Infection was associated with severity of cognitive impairment acutely and at 6-month follow-up independent of demographic factors and other confounders, including stroke severity. Infection was also associated with acute deficits in language, executive function, and attention domains, as well as 6-month deficits in number processing. Although inflammatory biomarkers including NLR, CRP and WCC were frequently elevated on admission, none were associated with cognitive impairment.

To our knowledge, there are only two previous studies of infection and cognitive outcome in stroke cohorts. One hospital-based study found an increased dementia risk in those with “hypoxic-ischaemic episodes”, defined as composite of “secondary insults”, including cardiac and thromboembolic events and infection, however numbers with infection were too small (pneumonia=2, sepsis=1) to allow separate analyses.^9^ The subsequent “Big Data” population-based study^8^ of administrative primary care data (>60,000 stroke survivors) found early dementia (3-12 months post-stroke) risk was elevated particularly after hospitalization-associated infection, which also increased the risk of late dementia.^6, 37^ However, the study design was retrospective and did not allow adjustment for stroke severity. Our study extends these previous findings by ascertaining cognitive impairment through prospective participant assessment, measuring infection severity directly using SIRS, and performing robust adjustment for a number of covariates.

Existing research suggests a biphasic inflammatory response to stroke, where an early activation phase is followed by systemic immunosuppression,^38, 39^ which increases susceptibility to infection.^38^ The prevalence of infection in our study (35% occurring on average around 4 days post-stroke) is consistent with the pooled prevalence in meta-analyses (30%) which also report older age, more severe stroke, dependency, and longer hospital admissions as risk factors.^10, 40^ We also recorded more pneumonia in dysphagic and NG-fed patients,^40^ and UTI in older, female patients, which is consistent with previous studies.^41^ Infection may also precipitate stroke; stroke risk increases immediately following onset of acute respiratory infections or UTI,^42^ and we cannot exclude the possibility that some infections were present pre-stroke, particularly since a fifth were identified on day of admission, of which a third were SIRS-positive.

Previous studies have shown that WCC, CRP and NLR are elevated in acute stroke, particularly with greater stroke severity, larger lesion volume, poor functional outcome, and recurrent stroke,^43, 44^ and the few data available suggest that similar factors are associated with the presence of SIRS.^45, 46^ Reported prevalence of SIRS post-stroke ranges from 14-36% on admission^45–49^ to 22-53% during admission,^49^ similar to the rates observed in our study (27% on admission, 45% during admission). Elevated inflammatory markers in acute stroke may also occur from chronic comorbidities, such as diabetes, hypertension, and smoking,^50^ though in our study smoking and diabetes rates were relatively uncommon. Comorbid load may also enhance the risk of SIRS,^51^ however the CCI score in our sample was relatively low.

Despite the prevalence of elevated inflammatory markers, we found no associations between CRP, WCC, or NLR and severity of cognitive impairment acutely or at 6 months. There are few previous studies on routinely available inflammatory biomarkers and post-stroke cognition, and findings are conflicting. Both positive and null associations with PSCI acutely and at 6-months have been reported for CRP^18–19^ and WCC.^52, 53^ Elevated NLR has been shown to be associated with global impairment at 3-months and visuospatial and memory domain impairment (NLR≥3.80),^17^ and executive dysfunction (MoCA) (NLR≥4.05).^54^ However, the sample characteristics and type/timing of cognitive assessment differed between studies. Taken together with our findings, the relationship between these individual inflammatory markers and post-stroke cognition remains uncertain. A recent study of an extensive panel of inflammatory biomarkers obtained using a research protocol (including TCC, interleukins, MIP-1α and TNF) found associations with baseline (but not chronic) biomarkers and cognitive impairment at up to 36 months’ follow-up suggesting that more sensitive/specific measures may detect inflammation relevant to cognitive outcomes.

We found SIRS (a composite measure of the inflammatory response using routinely measured markers) was linked to global cognitive impairment acutely, but not at follow-up in line with a previous study of SIRS and functional outcome at 3-6 months after stroke.^45^

Regarding specific cognitive domains, infection was associated with impairments in language, attention, and executive function acutely, and number processing at 6 months after adjustment for confounders. The pattern of impairments is consistent with that generally seen in cerebrovascular/small vessel disease (SVD) with relatively prominent deficits in frontal/executive domains.^24, 55, 56^ However, deficits particularly in attention may be seen in delirium, which may co-occur with infection and could have contributed to the acute findings although the requirement for informed consent will have excluded many/most with delirium.^57^ The lower number of frontal/executive domain associations at 6 months could have resulted from a reduction in power, as fewer individuals showed impairments across all domains at this time-point. The lack of a relationship with memory deficits is consistent with a recent UK Biobank study of infection and visual/verbal memory function although it should be noted that our study may have been underpowered.^7^ Our findings are also supported by a recent multi-cohort study^56^, in which hospitalization-associated infection was linked to a greater risk of vascular dementia vs Alzheimer’s disease suggesting greater cerebral susceptibility to infection in those with cerebrovascular disease.

Our study has some strengths. We addressed a key knowledge gap on the association of infection and routinely available inflammatory biomarkers with cognitive function in a representative sample of acute stroke patients (average age=74 years, 56% with NIHSS ≥5, 29% aphasic). We used a longitudinal study design and stroke-specific cognitive screen, with careful ascertainment of infection, inflammation biomarkers and measures of infection/inflammation severity through linkage to detailed hospital EPR data, supplemented by hand-searching of patient records. Limitations include that inflammatory biomarkers and SIRS data were not available for each day of admission as these were acquired as part of routine care, and under-ascertainment of peak values is possible, although acute stroke patients are usually comprehensively assessed and monitored. Finally, we could not exclude patients with pre-stroke infection or cognitive decline and therefore some of the association between infection and cognition could be driven by existing infection or increased susceptibility to infection in those with undiagnosed pre-stroke dementia. However, the requirement for informed consent means that most patients with dementia will likely have been excluded.

Our findings require replication in larger cohorts but have potential implications for the prevention of post-stroke dementia. Infection is thought to increase the risk of dementia through inflammatory mediators acting on the brain.^6, 7, 58^ Stroke patients may be particularly vulnerable to the effects of infection because of increased permeability of the blood brain barrier and co-existent small vessel disease.^58^ The pattern of cognitive domain impairment linked to infection is suggestive of a vascular profile and supports the findings from non-stroke studies of a preferential risk of vascular vs Alzheimer’s dementia after infection. Infection prevention and rapid treatment of infection may therefore help reduce dementia risk after stroke and likely explains some of the effect of organized stroke unit care on improved outcomes.^12, 45^ It may also present a tractable target for trials.

## Conclusions

Infection is prevalent in hospital stroke patients and is associated with cognitive impairment acutely and at 6-months follow-up with stronger associations for pneumonia and severe infection. In contrast, despite the frequent elevation in routinely measured inflammatory biomarkers (CRP, WCC, and NLR) indicating an acute inflammatory response, individual markers were not associated with cognitive impairment. Future research should aim to determine the underlying mechanisms linking infection to PSCI including the use of detailed inflammatory biomarker panels, and the relationship to cognitive outcomes in the longer term.

## Data Availability

All data produced in the present study are available upon reasonable request to the authors.

## Acknowledgements

The OCS studies were supported by the National Institute for Health Research (NIHR) Clinical Research Network. The views expressed are those of the authors and not necessarily those of the NHS, the NIHR or the Department of Health. We would like to thank all stroke survivors who participated, and members of the Oxford Translational Neuropsychology Group for contributions to recruitment, cognitive testing, and data entry.

