## Supplemental Material for "Infection, inflammation and post-stroke cognitive impairment"

### SUPPLEMENTARY MATERIALS

**Table S1a.** Charlson Comorbidity Index (CCI) scoring

| | CHARLSON COMORBIDITY INDEX (CCI) | |
|---|---|---|
| **Condition number** | **Condition description** | **Points** |
| 1 | Myocardial infarction | 1 |
| 2 | Congestive heart failure | 1 |
| 3 | Peripheral vascular disease | 1 |
| 4 | Cerebrovascular disease | 1 |
| 5 | Dementia | 1 |
| 6 | Chronic pulmonary disease | 1 |
| 7 | Rheumatic disease | 1 |
| 8 | Peptic ulcer disease | 1 |
| 9 | Liver disease, mild | 1 |
| 10 | Diabetes without chronic complications | 1 |
| 11 | Renal disease, mild to moderate | 1 |
| 12 | Diabetes with chronic complications | 2 |
| 13 | Hemiplegia or paraplegia | 2 |
| 14 | Any malignancy | 2 |
| 15 | Liver disease, moderate to severe | 3 |
| 16 | Renal disease, severe | 3 |
| 17 | HIV infection, no AIDS | 3 |
| 18 | Metastatic solid tumor | 6 |
| 19 | AIDS (HIV infection + opportunistic infection) | 6 |
| | *Maximum comorbidity score* | *29* |

**CDMF CCI Coding Schemes for all conditions (ICD-9 and ICD-10 diagnosis code):**
Glasheen WP, Cordier T, Gumpina R, Haugh G, Davis J, Renda A. Charlson Comorbidity Index: *ICD-9* Update and *ICD-10* Translation. Am Health Drug Benefits. 2019 Jun-Jul;12(4):188-197. PMID: 31428236

**Table S1b.** Charlson Comorbidity Index (CCI) scoring – Hierarchy rules.

| Category | Hierarchy Rule |
|---|---|
| 1 | Hemiplegia/paraplegia (Condition 13) trumps cerebrovascular disease (Condition 4) |
| 2 | Liver disease, moderate-severe (Condition 5) trumps liver disease, mild (Condition 9) |
| 3 | Diabetes with complications (Condition 12) trumps Diabetes, uncomplicated (Condition 10) |
| 4 | Renal disease, severe (Condition 16) trumps renal disease, mild-moderate (Condition 11) |
| 5 | Metastatic solid tumor (Condition 18) trumps malignancy (Condition 14) |
| 6 | AIDS (Condition 19) trumps HIV (Condition 14) |

AIDS, acquired immune deficiency syndrome; HIV, human immunodeficiency virus
N.B.: Within each hierarchy category, the milder condition should not contribute to the CCI score if the more severe condition applies even though codes for both may appear for an individual patient
N.B.: scoring supporting references:


1) Charlson ME, Pompei P, Ales KL, MacKenzie CR. A new method of classifying prognostic comorbidity in longitudinal studies: development and validation. J Chronic Dis. 1987;40(5):373-83. doi: 10.1016/0021-9681(87)90171-8.
2) Deyo RA, Cherkin DC, Ciol MA. Adapting a clinical comorbidity index for use with ICD-9-CM administrative databases. J Clin Epidemiol. 1992 Jun;45(6):613-9. doi: 10.1016/0895-4356(92)90133-8.
3) Quan H, Sundararajan V, Halfon P, Fong A, Burnand B, Luthi JC, Saunders LD, Beck CA, Feasby TE, Ghali WA. Coding algorithms for defining comorbidities in ICD-9-CM and ICD-10 administrative data. Med Care. 2005 Nov;43(11):1130-9. doi: 10.1097/01.mlr.0000182534.19832.83.

### Supplementary results

**Sensitivity analysis of missing acute NIHSS data:**
NIHSS data was available for n=215/255 (84.3%) of patients. To assess the potential impact of missing data, Little's MCAR Test was conducted to examine the randomness of missingness. The non-significant p-value (p=0.07) suggested that the missingness mechanism of the NIHSS data was likely random.

To address the missing NIHSS data, multiple imputation was performed using the 'mice' package in R. Sensitivity analyses were then conducted to compare the results between the imputed dataset (n=255) and a restricted dataset with complete NIHSS data (n=215). Notably, no significant differences were observed between the two sets of analyses. For instance, in both the adjusted models comprising complete cases and the imputed NIHSS dataset, the association between infection and post-stroke global cognitive impairment remained significant at follow-up. In the complete cases model (n=215), infection was associated with a significant increase in post-stroke cognitive impairment (β=5.23, SE=2.57, t=2.03, R2=0.204, p<0.05). Similarly, in the imputed NIHSS model (n=255), the association between infection and cognitive impairment remained significant (β=6.13, SE=2.47, t=2.49, R2=0.207, p<0.05).

**Table S2**. Proportion of cognitive impairments across all domains and subtests at both acute and 6-month follow-up (N = 255).

| Domain | Assessed acute (N) | Acute impaired (N) | % | Assessed follow-up (N) | Follow-up impaired (N) | % |
|---|---|---|---|---|---|---|
| **Language** | **255** | **110** | **43.14** | **255** | **79** | **30.98** |
| Picture naming | 255 | 78 | 30.59 | 255 | 46 | 18.04 |
| Semantic understanding | 255 | 28 | 10.98 | 255 | 11 | 4.31 |
| Sentence reading | 250 | 81 | 32.40 | 254 | 42 | 16.54 |
| **Attention** | **245** | **110** | **44.90** | **251** | **90** | **35.86** |
| Egocentric attention | 245 | 69 | 28.16 | 251 | 36 | 47.41 |
| Allocentric attention | 245 | 71 | 28.98 | 251 | 69 | 27.49 |
| **Executive Function** | **253** | **84** | **33.20** | **244** | **67** | **27.46** |
| **Memory** | **255** | **100** | **39.22** | **255** | **74** | **29.02** |
| Orientation | 255 | 54 | 21.18 | 254 | 48 | 18.90 |
| Verbal memory | 254 | 64 | 25.20 | 246 | 35 | 14.23 |
| Episodic memory | 254 | 49 | 19.29 | 246 | 20 | 8.13 |
| **Number processing** | **255** | **103** | **40.39** | **252** | **45** | **17.86** |
| Calculations | 255 | 36 | 14.12 | 252 | 9 | 3.57 |
| Writing | 254 | 97 | 38.19 | 252 | 43 | 17.06 |
| **Praxis** | **253** | **68** | **26.88** | **242** | **69** | **28.51** |
| Any domain | 255 | 213 | 83.53 | 255 | 192 | 75.29 |
| Single domain | 255 | 47 | 18.43 | 255 | 65 | 25.49 |
| Multi-domain | 255 | 166 | 65.10 | 255 | 127 | 49.80 |
| Unimpaired | 255 | 42 | 16.47 | 255 | 63 | 24.71 |

N.B.: The Oxford Cognitive Screen (OCS)[25] was designed as a short neuropsychological screen specifically for the stroke population with tasks which are inclusive for, and unconfounded by, aphasia, apraxia, and neglect. This study assessed 12 subtest scores covering 6 cognitive domains: language (picture naming, semantic understanding, sentence reading), attention (egocentric and sustained attention, allocentric attention), executive function (trailmaking), memory (orientation, verbal memory, episodic memory), praxis (gesture imitation), and number processing (calculation and number writing). Subtests were binarized into impaired or unimpaired based on normative scores for each subtest (cut-offs were set at 5th centile).[25] There is no single maximum score, as each subtest/task has a different threshold cut off to indicate impairment based on the published normative data. A domain impairment was characterized as at least one impaired subtest in that domain as number of subtests range from 1-3 across domains. The OCS takes approximately 15-20 minutes to administer and was administered by trained neuropsychologists and occupational therapists at both bedside acutely and at in-person follow-up assessments.

**Table S3.** Causative organisms found for Urinary tract infections (UTIs), soft tissue infections, and blood infections.

| | **N (%)** |
|---|---|
| **UTI** | 39 (15.29) |
| Citrobacter species | 2 |
| Escherichia coli | 13 |
| Enterobacter aerogenes | 1 |
| Enterobacter cloacae | 1 |
| Enterococcus faecalis | 3 |
| Enterococcus faecium | 1 |
| Klebsiella oxytoca | 1 |
| Klebsiella pneumoniae | 1 |
| Klebsiella species | 1 |
| Mixed growth | 10 |
| Proteus mirabilis | 1 |
| Proteus species | 1 |
| Staphylococcus epidermidis | 1 |
| Suspected | 1 |
| **Soft tissue** | 9 (3.53) |
| Candidiasis | 4 |
| MRSA | 3 |
| Pseudomonas aeruginosa | 1 |
| Serratia marcescens | 1 |
| **Blood** | 8 (3.14) |
| Escherichia coli | 1 |
| Coagulase-negative staphylococcus | 1 |
| Serratia Marcescens | 1 |
| Staphylococcus | 1 |
| Staphylococcus capitis | 1 |
| Staphylococcus epidermidis | 2 |
| Streptococcus agalactiae | 1 |

**Table S4**. Multivariate regression results of the association between SIRS on admission and severity of cognitive impairment acutely and at 6 months.

| | ACUTE | | | | 6 MONTHS | | | |
|---|---|---|---|---|---|---|---|---|
| *Adjusted models* | β | *SE* | *t* | *p* | β | *SE* | *t* | *p* |
| **SIRS (admission)** | 9.190 | 3.625 | 2.535 | **0.012** | 1.795 | 2.381 | 0.754 | 0.452 |
| *Covariates* | | | | | | | | |
| Age | 0.220 | 0.142 | 1.547 | 0.124 | 0.399 | 0.933 | 4.276 | **<0.0001** |
| Sex | 0.404 | 3.214 | 0.126 | 0.900 | 0.383 | 2.111 | 0.181 | 0.856 |
| Education | -0.715 | 0.433 | -1.653 | 0.100 | -0.829 | 0.284 | -2.918 | **0.004** |
| NIHSS | 1.242 | 0.281 | 4.412 | **<0.0001** | 0.0108 | 0.185 | 0.586 | 0.559 |
| Previous stroke | 3.600 | 3.465 | 1.039 | 0.300 | 0.198 | 2.277 | 0.087 | 0.931 |
| Diabetes | 7.447 | 4.117 | 1.809 | 0.072 | 0.912 | 2.705 | 0.337 | 0.736 |
| Hypertension | 3.499 | 4.094 | 0.855 | 0.394 | 0.889 | 2.195 | 0.405 | 0.686 |
| Atrial fibrillation | 2.511 | 3.914 | 0.641 | 0.522 | 0.956 | 2.572 | 0.372 | 0.711 |
| Smoking | 3.499 | 3.625 | 2.535 | 0.012 | 0.918 | 2.689 | 0.341 | 0.733 |

Severity of cognitive impairment was defined by proportion of Oxford Cognitive Screen tasks impaired.
Model $R^2$ acute = 0.176; 6-months = 0.186

**Table S5**. Multivariate regression results of the association between infection and severity of cognitive impairment acutely and at 6 months.

| | Severity of cognitive impairment | | | | | | | |
|---|---|---|---|---|---|---|---|---|
| | ACUTE | | | | 6 MONTHS | | | |
| *Adjusted models* | β | *SE* | *t* | *p* | β | *SE* | *t* | *p* |
| **Infection (any)** | 6.349 | 3.021 | 2.102 | **0.037** | 6.132 | 2.466 | 2.486 | **0.014** |
| *Covariates* | | | | | | | | |
| Age | 0.112 | 0.127 | 0.881 | 0.379 | 0.387 | 0.081 | 4.788 | **<0.0001** |
| Sex | 0.553 | 2.923 | 0.189 | 0.850 | -0.157 | 2.074 | -0.076 | 0.940 |
| Education | -0.801 | 0.408 | -1.963 | 0.051 | -0.977 | 0.254 | -3.847 | **0.0002** |
| NIHSS | 1.515 | 0.261 | 5.806 | **<0.0001** | 0.486 | 0.226 | 2.146 | **0.033** |
| Previous stroke | 2.856 | 3.192 | 0.895 | 0.372 | 0.472 | 2.197 | 0.215 | 0.830 |
| Diabetes | 7.662 | 3.773 | 2.031 | **0.043** | 0.291 | 2.903 | 0.100 | 0.920 |
| Hypertension | 4.407 | 3.037 | 1.451 | 0.148 | 1.112 | 2.167 | 0.513 | 0.608 |
| Atrial fibrillation | 2.774 | 3.460 | 0.802 | 0.424 | 0.978 | 2.368 | 0.413 | 0.680 |
| Smoking | 2.039 | 3.771 | 0.541 | 0.589 | 2.800 | 2.788 | 1.004 | 0.312 |

Severity of cognitive impairment was defined by proportion of Oxford Cognitive Screen tasks impaired.
Model $R^2$ acute = 0.188; 6 months = 0.207

**Table S6.** Multivariate regression results of the association between types of infection (pneumonia and UTI) and severity of cognitive impairment acutely and at 6 months.

| | **Severity of cognitive impairment** | | | | | | | |
|---|---|---|---|---|---|---|---|---|
| | **ACUTE** | | | | **6 MONTHS** | | | |
| *Adjusted models* | β | *SE* | *t* | *p* | β | *SE* | *t* | *p* |
| **Pneumonia** | 8.978 | 3.769 | 2.382 | **0.018** | 5.837 | 2.690 | 2.171 | **0.031** |
| **UTI** | 1.913 | 4.080 | 0.469 | 0.640 | 3.282 | 2.911 | 1.127 | 0.261 |
| *Covariates* | | | | | | | | |
| Age | 0.113 | 0.127 | 0.887 | 0.376 | 0.394 | 0.091 | 4.345 | **<0.0001** |
| Sex | 0.429 | 2.951 | 0.146 | 0.884 | -0.4334 | 2.106 | -0.206 | 0.837 |
| Education | -0.765 | 0.408 | -1.874 | 0.062 | -0.949 | 0.291 | -3.260 | **0.001** |
| NIHSS | 1.4401 | 0.264 | 5.450 | **<0.0001** | 0.446 | 0.189 | 2.368 | **0.019** |
| Previous stroke | 2.853 | 3.189 | 0.895 | 0.372 | 0.377 | 2.26 | -0.166 | 0.869 |
| Diabetes | 7.555 | 3.778 | 2.000 | **0.047** | 0.251 | 2.700 | 0.093 | 0.926 |
| Hypertension | 4.932 | 3.03 | 11.628 | 0.105 | 1.557 | 2.162 | 0.720 | 0.472 |
| Atrial fibrillation | 2.992 | 3.454 | 0.866 | 0.387 | 0.818 | 2.465 | 0.332 | 0.740 |
| Smoking | 1.926 | 3.786 | 0.509 | 0.611 | 2.963 | 2.702 | 1.097 | 0.274 |

Severity of cognitive impairment was defined by proportion of Oxford Cognitive Screen tasks impaired.
Model $R^2$ acute = 0.194; 6 months = 0.202

**Table S7.** Association between severity of infection determined by the presence or absence of SIRS anytime and severity of cognitive impairment acutely and at 6 months.

| | **Severity of cognitive impairment** | | | | | | | |
|---|---|---|---|---|---|---|---|---|
| | **ACUTE** | | | | **6 MONTHS** | | | |
| *Adjusted models* | β | *SE* | *t* | *p* | β | *SE* | *t* | *p* |
| **Infection, SIRS-negative** | 4.112 | 4.991 | 0.824 | 0.411 | 3.92 | 4.238 | 0.800 | 0.425 |
| **Infection, SIRS-positive** | 8.840 | 3.851 | 2.296 | **0.023** | 5.085 | 2.883 | 1.764 | 0.079 |
| *Covariates* | | | | | | | | |
| Age | 0.158 | 0.144 | 1.094 | 0.275 | 0.434 | 0.094 | 4.615 | **<0.0001** |
| Sex | 0.647 | 3.236 | 0.200 | 0.842 | 0.255 | 2.220 | 0.115 | 0.909 |
| Education | -0.662 | 0.436 | -1.521 | 0.130 | -0.948 | 0.258 | -3.680 | **0.0003** |
| NIHSS | 1.232 | 0.284 | 4.337 | **<0.0001** | 0.185 | 0.212 | 0.873 | 0.384 |
| Previous stroke | 4.592 | 3.509 | 1.309 | 0.192 | 1.051 | 2.374 | 0.443 | 0.659 |
| Diabetes | 7.133 | 4.146 | 1.721 | 0.087 | 0.036 | 3.294 | 0.321 | 0.784 |
| Hypertension | 4.606 | 3.390 | 1.359 | 0.176 | 0.602 | 2.334 | 0.258 | 0.797 |
| Atrial fibrillation | 0.858 | 3.863 | 0.222 | 0.825 | 0.207 | 2.640 | 0.078 | 0.937 |
| Smoking | 2.620 | 4.146 | 1.721 | 0.087 | 0.955 | 2.973 | 0.321 | 0.748 |

Severity of cognitive impairment was defined by proportion of Oxford Cognitive Screen tasks impaired.
Model $R^2$ acute = 0.172, 6 months = 0.233

**Table S8.** Associations between acute post-stroke infection and domain-specific cognitive impairments acutely and at 6 months.

| *Adjusted models* | | | | | | | |
|---|---|---|---|---|---|---|---|
| **ACUTE** | β | *SE* | *z* | *p* | $R^2$ | OR | 95% CI |
| Language | 0.545 | 0.277 | 1.967 | **0.049** | 0.065 | 1.725 | 1.002 - 2.971 |
| Attention | 0.696 | 0.287 | 2.422 | **0.015** | 0.106 | 2.006 | 1.142 - 3.522 |
| Executive function | 0.632 | 0.297 | 2.125 | **0.034** | 0.114 | 1.881 | 1.050 - 3.367 |
| Memory | 0.474 | 0.281 | 1.658 | 0.092 | 0.071 | 1.606 | 0.926 - 2.785 |
| Number processing | 0.273 | 0.285 | 0.957 | 0.338 | 0.089 | 1.314 | 0.751 - 2.297 |
| Praxis | 0.156 | 0.304 | 0.512 | 0.609 | 0.038 | 1.169 | 0.643 - 2.123 |
| **6 MONTHS** | | | | | | | |
| Language | 0.389 | 0.295 | 1.317 | 0.188 | 0.086 | 1.475 | 0.827 - 2.631 |
| Attention | 0.528 | 0.287 | 1.842 | 0.066 | 0.010 | 1.695 | 0.967 - 2.972 |
| Executive function | 0.359 | 0.314 | 1.145 | 0.252 | 0.100 | 1.433 | 0.774 - 2.651 |
| Memory | 0.565 | 0.301 | 1.880 | 0.060 | 0.100 | 1.760 | 0.976 - 3.174 |
| Number processing | 0.759 | 0.351 | 2.162 | **0.031** | 0.106 | 2.137 | 1.073 - 4.254 |
| Praxis | 0.024 | 0.321 | 0.075 | 0.941 | 0.090 | 1.024 | 0.546 - 1.920 |

Each Oxford Cognitive Screen (OCS) domain was binarized into impaired/unimpaired. Adjusted models covariates included age, sex, education, stroke severity, previous stroke, atrial fibrillation, hypertension, diabetes mellitus, and smoking. $R^2$ refers to McFadden's pseudo $R^2$ for each model; OR: odds ratio (log scale).

**Figure S1**. Other than infection, significant correlates of domain-specific impairments within each adjusted model (demographic and vascular covariates) included:

- **Language**
  - *Acute* – education (*p* <0.01), stroke severity (*p* <0.05)
  - *6 months* – age (*p* <0.05) and education (*p* <0.01)
- **Attention**
  - *Acute* – stroke severity (*p* <0.001), atrial fibrillation (AF) (*p* <0.05), diabetes (DM) (*p* <0.01)
  - *6 months* – age (*p* <0.001) and education (*p* <0.01)
- **Executive function**
  - *Acute* – stroke severity (*p* <0.001) and DM (*p* <0.05)
  - *6 months* – age (*p* <0.01), Sex (female) *p* <0.05), stroke severity (*p* <0.05)
- **Memory**
  - *Acute* – stroke severity *p* <0.001) and DM (*p* <0.05)
  - *6 months* – age (*p* <0.05), education (*p* <0.06), smoking (*p* <0.05)
- **Number processing**
  - *Acute* – education (*p* <0.001), stroke severity (*p* <0.01)
  - *6 months* – education (*p* <0.05) and hypertension (*p* <0.05)
- **Praxis**
  - *Acute* – hypertension (*p* <0.05)
  - *6 months* – age (*p* <0.01), sex (male) (*p* <0.001)
